# Quality of care at safety-net hospitals and the impact on pay-for-performance reimbursement

**DOI:** 10.1101/19010843

**Authors:** Reith R. Sarkar, Patrick T. Courtney, Katie Bachand, Paige Sheridan, Paul Riviere, Zachary D. Guss, Christian R. Lopez, Michael G. Brandel, Matthew P. Banegas, James D. Murphy

**Author notes:** **Correspondence to:** James D. Murphy, M.D., M.S., University of California, San Diego, Altman Clinical and Translational Research Institute, 9452 Medical Center Dr, La Jolla, CA 92037, / /. **Support:** NIH TL1 TR001443 (RRS, PTC, PR, MGB), NIH U54 CA132379 (JDM). **Disclosures:** RRS and JDM receive compensation for consulting from Boston Consulting Group. PR receives compensation for consulting from Peptide Logic, LLC.

## Abstract

**Background:** Pay-for-performance reimbursement ties hospital payments to standardized quality of care metrics. The impact of pay-for-performance reimbursement models on safety-net hospitals, which care primarily for uninsured or underinsured patients, remains poorly defined. This study evaluates how standardized quality of care metrics vary by a hospital’s safety-net status, and helps us better understand the potential impact that pay-for-performance reimbursement could have on funding of safety-net hospitals.

**Methods:** We identified 1,703,865 bladder, breast, cervix, colon, endometrium, gastric, lung, ovary, or rectum cancer patients treated at 1,344 hospitals diagnosed between 2004 and 2015. Safety-net burden was defined for each hospital as the percentage of uninsured or Medicaid patients cared for by that hospital. Hospitals were grouped into low-, medium-, and high-burden hospitals. We evaluated the impact of safety-net burden on concordance with 20 standardized quality of care measures, adjusting for differences in patient age, gender, stage at diagnosis, and comorbidity.

**Results:** Patients seen at high-burden hospitals were more likely to be young, male, black, Hispanic, and to reside in a low-income and low-educated region. High-burden hospitals had lower adherence to 13 of 20 quality measures compared to low-burden hospitals (all p<0.05). Among the 350 high-burden hospitals, the quality measures were lowest for those caring for the highest fraction of uninsured or Medicaid patients, minority serving hospitals, and those caring for less educated patients (all p<0.001).

**Discussion:** Cancer care at safety-net hospitals was associated with lower concordance to standardized quality of care measures. Under a pay-for-performance reimbursement model these lower quality of care scores could decrease payments to safety-net hospitals, potentially increasing health disparities for at-risk cancer patients.

## Background

Reimbursement for healthcare in the United States (US) has steadily moved from a fee-for-service reimbursement model to pay-for-performance or value-based reimbursement models^1-3^. Paying providers and hospitals for high quality care (pay-for-performance), as opposed to simply paying for providing care (fee-for-service), appeals to multiple stakeholders. Modern pay-for-performance reimbursement strategies depend on standardized assessments of quality of care, and typically hospitals with higher aggregate quality scores receive performance bonuses, whereas hospitals performing poorly incur financial penalties. In general, the quality of care metrics used to define reimbursement are adjusted to account for differences in patient age, gender, geography, disease severity, and underlying patient comorbidity. However, payers such as Medicare typically do not account for differences in race, ethnicity, socioeconomic status, or other social determinants of health, based on the argument that hospitals with higher percentages of underserved patients should not be held to a lower standard of care^4^.

Safety-net hospitals provide care for a large proportion of uninsured and Medicaid-insured patients, and overall provide a valuable service for underserved individuals across the US^5-7^. Given the insurance status of patients cared for by safety-net hospitals, these institutions at baseline receive lower reimbursement for providing healthcare, and therefore often struggle financially^8^. Furthermore, the population of patients treated by safety-net hospitals often face social, economic, and logistic barriers to receiving high quality timely healthcare^9-11^. Under a pay-for-performance reimbursement model, these barriers to providing high quality care could reduce hospital revenue for safety-net-hospitals, which could indirectly widen health disparities among an at-risk population of patients. In areas outside of oncology, research demonstrates that pay-for-performance reimbursement policies could lead to decreased payments to hospitals caring for underserved patients^12,13^, though this question has not been addressed within the field of oncology. We hypothesize that the complex and multidisciplinary treatment required for timely, high quality cancer care could lead to decreased quality of cancer care at safety-net hospitals. This in turn could lead to decreased reimbursement under a pay-for-performance reimbursement model. The purpose of this study was to evaluate the relationship between safety-net hospitals and standard quality of care metrics in a large population-based cohort of cancer patients. Understanding this relationship will provide insight into the impact that pay-for-performance reimbursement for oncology could have on safety-net hospitals.

## Methods

### Data source

Patients were identified from the National Cancer Database (NCDB), which is a large cancer registry jointly supported by the American College of Surgeons and the American Cancer Society. The NCDB represents a hospital-based cancer registry that collects patient clinical and treatment data on incident cancer cases diagnosed from over 1,500 facilities, which covers 70% of cancer cases diagnosed each year in the United States.

### Quality of care measures

The Commission on Cancer (CoC) is a consortium of professional organizations dedicated to improving survival and quality of life for cancer patients through standard-setting, prevention, research, education, and the monitoring of comprehensive quality care^14^. The CoC has produced several cancer quality of care measures which directly draw from data collected within cancer registries^15^. Each quality measure focuses on one of three areas: accountability, quality improvement, or surveillance. *Accountability* measures are backed by high levels of evidence typically including multiple randomized trials. *Quality improvement* measures are not backed by randomized trials, and are derived from lower levels of non-randomized evidence. *Surveillance* measures have more limited evidence, and often reflect the status quo. We evaluated 20 quality of care measures across nine cancer sites: bladder, breast, cervix, colon, endometrium, gastric, non-small cell lung, ovary, and rectum. Descriptions of the 20 quality of care measures included in this study are included in **Table 1**.

**Table 1.**
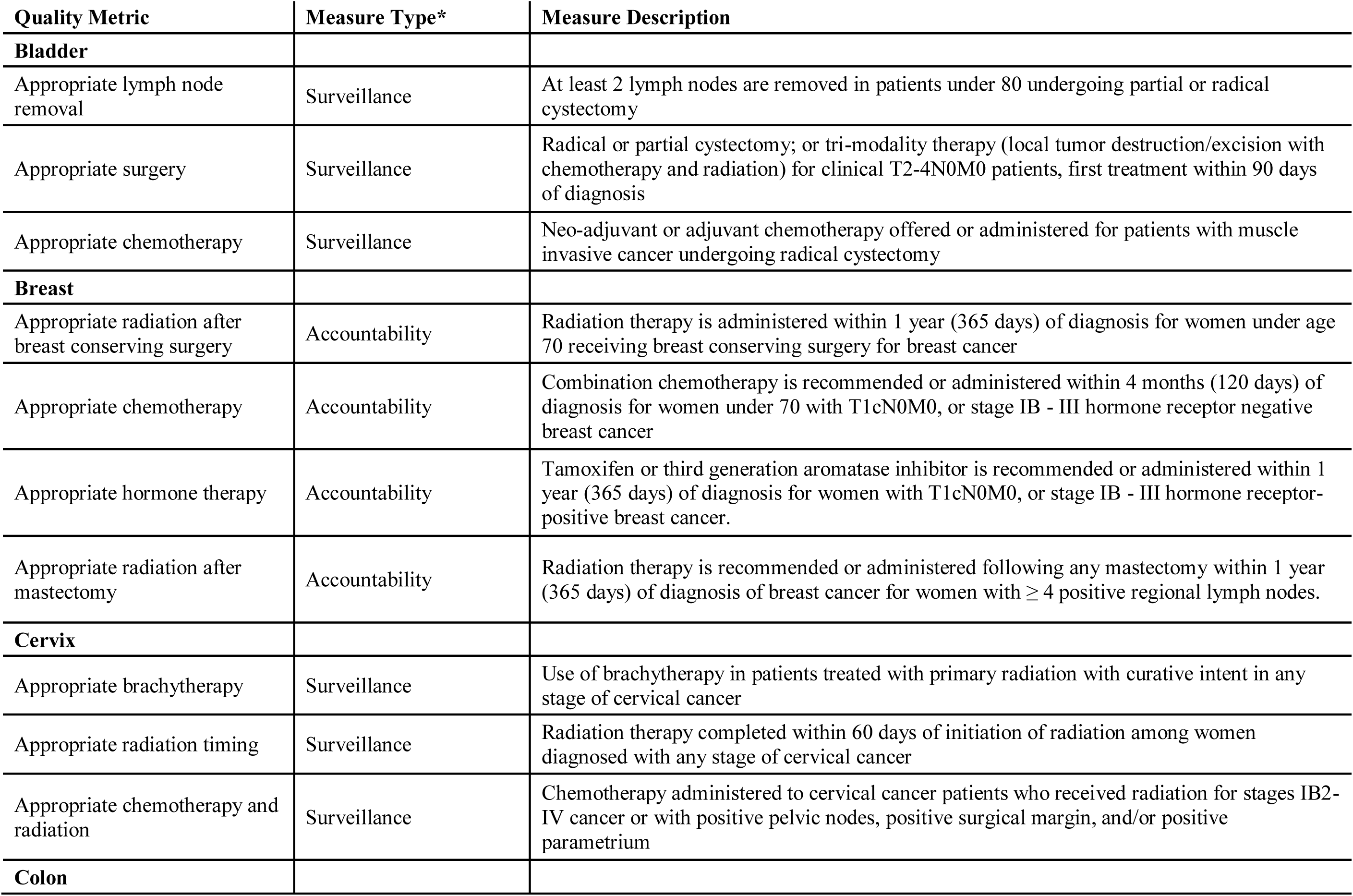
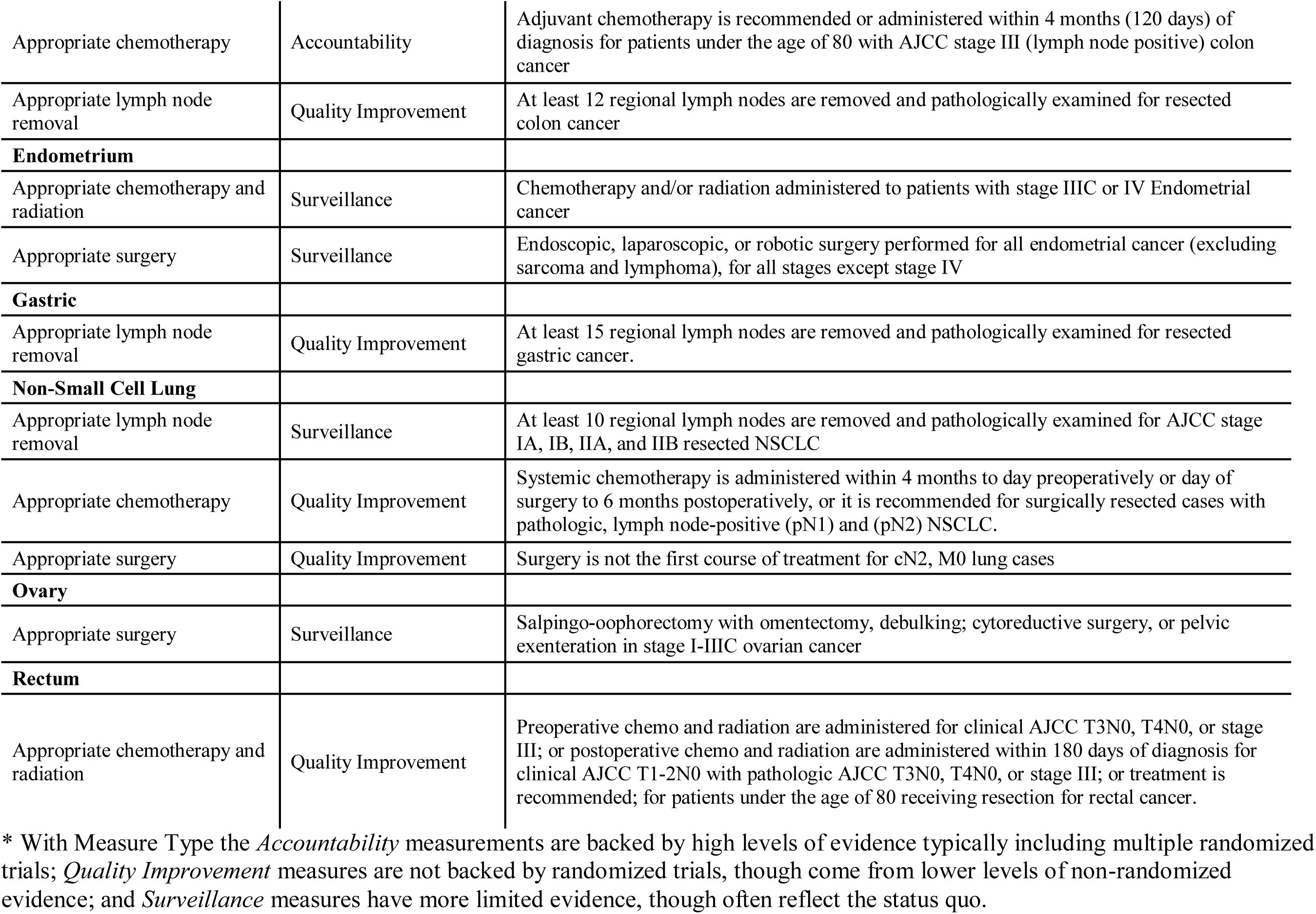
Definition of Quality Measures.

| **Quality Metric** | **Measure Type\*** | **Measure Description** |
| --- | --- | --- |
| **Bladder** |  |  |
| Appropriate lymph node removal | Surveillance | At least 2 lymph nodes are removed in patients under 80 undergoing partial or radical cystectomy |
| Appropriate surgery | Surveillance | Radical or partial cystectomy; or tri-modality therapy (local tumor destruction/excision with chemotherapy and radiation) for clinical T2-4N0M0 patients, first treatment within 90 days of diagnosis |
| Appropriate chemotherapy | Surveillance | Neo-adjuvant or adjuvant chemotherapy offered or administered for patients with muscle invasive cancer undergoing radical cystectomy |
| **Breast** |  |  |
| Appropriate radiation after breast conserving surgery | Accountability | Radiation therapy is administered within 1 year (365 days) of diagnosis for women under age 70 receiving breast conserving surgery for breast cancer |
| Appropriate chemotherapy | Accountability | Combination chemotherapy is recommended or administered within 4 months (120 days) of diagnosis for women under 70 with T1cN0M0, or stage IB - III hormone receptor negative breast cancer |
| Appropriate hormone therapy | Accountability | Tamoxifen or third generation aromatase inhibitor is recommended or administered within 1 year (365 days) of diagnosis for women with T1cN0M0, or stage IB - III hormone receptor-positive breast cancer. |
| Appropriate radiation after mastectomy | Accountability | Radiation therapy is recommended or administered following any mastectomy within 1 year (365 days) of diagnosis of breast cancer for women with $\geq 4$ positive regional lymph nodes. |
| **Cervix** |  |  |
| Appropriate brachytherapy | Surveillance | Use of brachytherapy in patients treated with primary radiation with curative intent in any stage of cervical cancer |
| Appropriate radiation timing | Surveillance | Radiation therapy completed within 60 days of initiation of radiation among women diagnosed with any stage of cervical cancer |
| Appropriate chemotherapy and radiation | Surveillance | Chemotherapy administered to cervical cancer patients who received radiation for stages IB2-IV cancer or with positive pelvic nodes, positive surgical margin, and/or positive parametrium |
| **Colon** |  |  |
| Appropriate chemotherapy | Accountability | Adjuvant chemotherapy is recommended or administered within 4 months (120 days) of diagnosis for patients under the age of 80 with AJCC stage III (lymph node positive) colon cancer |
| Appropriate lymph node removal | Quality Improvement | At least 12 regional lymph nodes are removed and pathologically examined for resected colon cancer |
| **Endometrium** |  |  |
| Appropriate chemotherapy and radiation | Surveillance | Chemotherapy and/or radiation administered to patients with stage IIIC or IV Endometrial cancer |
| Appropriate surgery | Surveillance | Endoscopic, laparoscopic, or robotic surgery performed for all endometrial cancer (excluding sarcoma and lymphoma), for all stages except stage IV |
| **Gastric** |  |  |
| Appropriate lymph node removal | Quality Improvement | At least 15 regional lymph nodes are removed and pathologically examined for resected gastric cancer. |
| **Non-Small Cell Lung** |  |  |
| Appropriate lymph node removal | Surveillance | At least 10 regional lymph nodes are removed and pathologically examined for AJCC stage IA, IB, IIA, and IIB resected NSCLC |
| Appropriate chemotherapy | Quality Improvement | Systemic chemotherapy is administered within 4 months to day preoperatively or day of surgery to 6 months postoperatively, or it is recommended for surgically resected cases with pathologic, lymph node-positive (pN1) and (pN2) NSCLC. |
| Appropriate surgery | Quality Improvement | Surgery is not the first course of treatment for cN2, M0 lung cases |
| **Ovary** |  |  |
| Appropriate surgery | Surveillance | Salpingo-oophorectomy with omentectomy, debulking; cytoreductive surgery, or pelvic exenteration in stage I-IIIC ovarian cancer |
| **Rectum** |  |  |
| Appropriate chemotherapy and radiation | Quality Improvement | Preoperative chemo and radiation are administered for clinical AJCC T3N0, T4N0, or stage III; or postoperative chemo and radiation are administered within 180 days of diagnosis for clinical AJCC T1-2N0 with pathologic AJCC T3N0, T4N0, or stage III; or treatment is recommended; for patients under the age of 80 receiving resection for rectal cancer. |

\* With Measure Type the *Accountability* measurements are backed by high levels of evidence typically including multiple randomized trials; *Quality Improvement* measures are not backed by randomized trials, though come from lower levels of non-randomized evidence; and *Surveillance* measures have more limited evidence, though often reflect the status quo.

### Patient selection

The individual quality of care measures inherently assess quality among patients with different tumor sites, stages and treatments. Our initial patient selection strategy within NCDB identified patients using standard inclusion criteria to identify eligible patients for each quality metric^16^. This initial query identified 1,762,441 patients aged 18 and over with the appropriate stage and treatment diagnosed between 2004 and 2015. Our estimation of hospital safety-net status required knowledge of insurance status, therefore our initial query only included subjects with known insurance status. We excluded 58,576 patients with missing tumor- or treatment-related variables required to calculate specific quality of care endpoints. **Supplementary Table 1** demonstrates details of the patient selection process. There was no difference in the proportions of Medicaid and uninsured patients versus insured patients included in the initial query compared to the final study cohort. The final study cohort included 1,703,865 patients (**Table 2**).

**Table 2.**
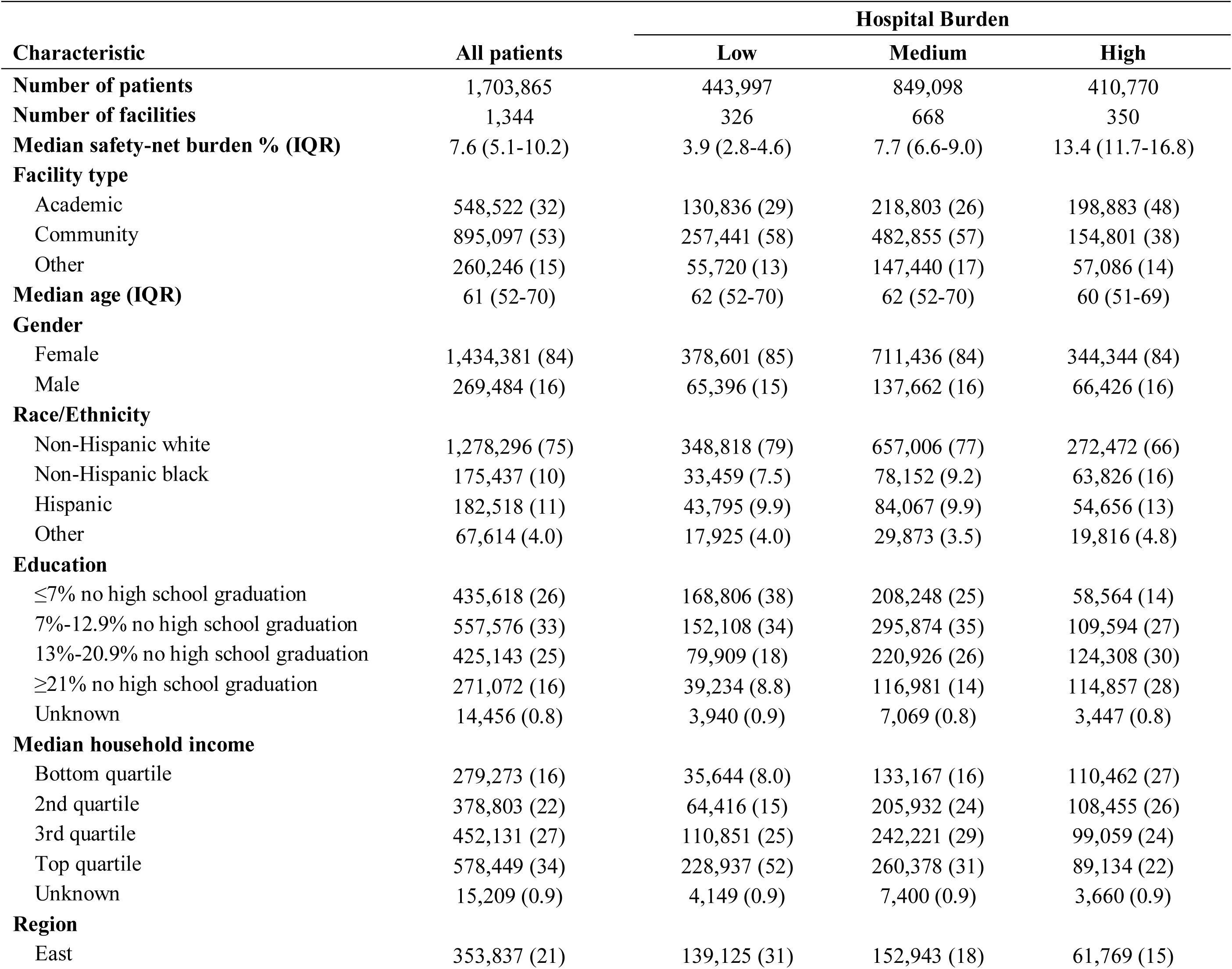
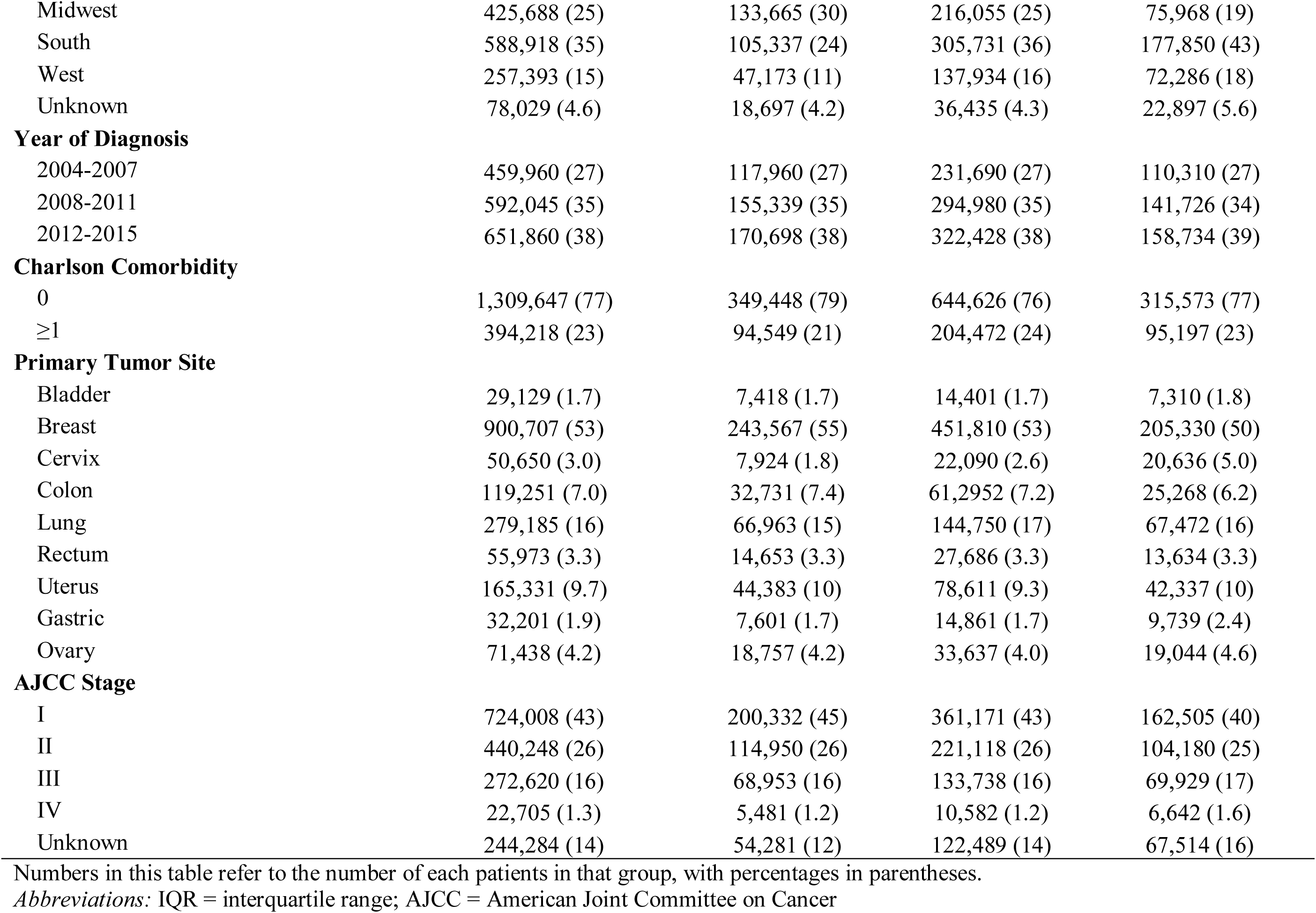
Patient Characteristics.

| Characteristic | All patients | Hospital Burden |  |  |
| --- | --- | --- | --- | --- |
|  |  | Low | Medium | High |
| **Number of patients** | 1,703,865 | 443,997 | 849,098 | 410,770 |
| **Number of facilities** | 1,344 | 326 | 668 | 350 |
| **Median safety-net burden % (IQR)** | 7.6 (5.1-10.2) | 3.9 (2.8-4.6) | 7.7 (6.6-9.0) | 13.4 (11.7-16.8) |
| **Facility type** |  |  |  |  |
| Academic | 548,522 (32) | 130,836 (29) | 218,803 (26) | 198,883 (48) |
| Community | 895,097 (53) | 257,441 (58) | 482,855 (57) | 154,801 (38) |
| Other | 260,246 (15) | 55,720 (13) | 147,440 (17) | 57,086 (14) |
| **Median age (IQR)** | 61 (52-70) | 62 (52-70) | 62 (52-70) | 60 (51-69) |
| **Gender** |  |  |  |  |
| Female | 1,434,381 (84) | 378,601 (85) | 711,436 (84) | 344,344 (84) |
| Male | 269,484 (16) | 65,396 (15) | 137,662 (16) | 66,426 (16) |
| **Race/Ethnicity** |  |  |  |  |
| Non-Hispanic white | 1,278,296 (75) | 348,818 (79) | 657,006 (77) | 272,472 (66) |
| Non-Hispanic black | 175,437 (10) | 33,459 (7.5) | 78,152 (9.2) | 63,826 (16) |
| Hispanic | 182,518 (11) | 43,795 (9.9) | 84,067 (9.9) | 54,656 (13) |
| Other | 67,614 (4.0) | 17,925 (4.0) | 29,873 (3.5) | 19,816 (4.8) |
| **Education** |  |  |  |  |
| ≤7% no high school graduation | 435,618 (26) | 168,806 (38) | 208,248 (25) | 58,564 (14) |
| 7%-12.9% no high school graduation | 557,576 (33) | 152,108 (34) | 295,874 (35) | 109,594 (27) |
| 13%-20.9% no high school graduation | 425,143 (25) | 79,909 (18) | 220,926 (26) | 124,308 (30) |
| ≥21% no high school graduation | 271,072 (16) | 39,234 (8.8) | 116,981 (14) | 114,857 (28) |
| Unknown | 14,456 (0.8) | 3,940 (0.9) | 7,069 (0.8) | 3,447 (0.8) |
| **Median household income** |  |  |  |  |
| Bottom quartile | 279,273 (16) | 35,644 (8.0) | 133,167 (16) | 110,462 (27) |
| 2nd quartile | 378,803 (22) | 64,416 (15) | 205,932 (24) | 108,455 (26) |
| 3rd quartile | 452,131 (27) | 110,851 (25) | 242,221 (29) | 99,059 (24) |
| Top quartile | 578,449 (34) | 228,937 (52) | 260,378 (31) | 89,134 (22) |
| Unknown | 15,209 (0.9) | 4,149 (0.9) | 7,400 (0.9) | 3,660 (0.9) |
| **Region** |  |  |  |  |
| East | 353,837 (21) | 139,125 (31) | 152,943 (18) | 61,769 (15) |
| Midwest | 425,688 (25) | 133,665 (30) | 216,055 (25) | 75,968 (19) |
| South | 588,918 (35) | 105,337 (24) | 305,731 (36) | 177,850 (43) |
| West | 257,393 (15) | 47,173 (11) | 137,934 (16) | 72,286 (18) |
| Unknown | 78,029 (4.6) | 18,697 (4.2) | 36,435 (4.3) | 22,897 (5.6) |
| **Year of Diagnosis** |  |  |  |  |
| 2004-2007 | 459,960 (27) | 117,960 (27) | 231,690 (27) | 110,310 (27) |
| 2008-2011 | 592,045 (35) | 155,339 (35) | 294,980 (35) | 141,726 (34) |
| 2012-2015 | 651,860 (38) | 170,698 (38) | 322,428 (38) | 158,734 (39) |
| **Charlson Comorbidity** |  |  |  |  |
| 0 | 1,309,647 (77) | 349,448 (79) | 644,626 (76) | 315,573 (77) |
| ≥1 | 394,218 (23) | 94,549 (21) | 204,472 (24) | 95,197 (23) |
| **Primary Tumor Site** |  |  |  |  |
| Bladder | 29,129 (1.7) | 7,418 (1.7) | 14,401 (1.7) | 7,310 (1.8) |
| Breast | 900,707 (53) | 243,567 (55) | 451,810 (53) | 205,330 (50) |
| Cervix | 50,650 (3.0) | 7,924 (1.8) | 22,090 (2.6) | 20,636 (5.0) |
| Colon | 119,251 (7.0) | 32,731 (7.4) | 61,2952 (7.2) | 25,268 (6.2) |
| Lung | 279,185 (16) | 66,963 (15) | 144,750 (17) | 67,472 (16) |
| Rectum | 55,973 (3.3) | 14,653 (3.3) | 27,686 (3.3) | 13,634 (3.3) |
| Uterus | 165,331 (9.7) | 44,383 (10) | 78,611 (9.3) | 42,337 (10) |
| Gastric | 32,201 (1.9) | 7,601 (1.7) | 14,861 (1.7) | 9,739 (2.4) |
| Ovary | 71,438 (4.2) | 18,757 (4.2) | 33,637 (4.0) | 19,044 (4.6) |
| **AJCC Stage** |  |  |  |  |
| I | 724,008 (43) | 200,332 (45) | 361,171 (43) | 162,505 (40) |
| II | 440,248 (26) | 114,950 (26) | 221,118 (26) | 104,180 (25) |
| III | 272,620 (16) | 68,953 (16) | 133,738 (16) | 69,929 (17) |
| IV | 22,705 (1.3) | 5,481 (1.2) | 10,582 (1.2) | 6,642 (1.6) |
| Unknown | 244,284 (14) | 54,281 (12) | 122,489 (14) | 67,514 (16) |

Numbers in this table refer to the number of each patients in that group, with percentages in parentheses.
*Abbreviations:* IQR = interquartile range; AJCC = American Joint Committee on Cancer

### Hospital safety-net burden

As defined by the Institute of Medicine, healthcare safety-net hospitals refer to facilities that deliver a significant level of healthcare to patients with no insurance or Medicaid^17^. We defined each hospital’s safety-net burden as the proportion of cancer patients cared for throughout the study period without insurance or with Medicaid. To measure safety-net burden, we used all solid tumor cancer patients within NCDB diagnosed between 2004 and 2015 with known insurance status (4,696,779 patients), as opposed to using only patients in this study. Similar to prior research, we divided hospitals by safety-net burden into quartiles and assigned the lowest quartile as *low-burden hospitals*, the middle 2 quartiles as *medium-burden hospitals*, and the highest quartile as *high-burden hospitals*^18^. Therefore, high burden hospitals care for the largest fraction of uninsured or Medicaid patients.

### Study covariables

Study covariables included patient factors such as age at diagnosis, year of diagnosis, gender, race, ethnicity, and Charlson comorbidity score. Socioeconomic information included zip-code level median household income and high school graduation rate. Clinical factors included AJCC 6/7^th^ edition tumor stage.

### Statistical analysis

Pearson chi-square tests and Wilcoxon rank sum tests were used to identify differences in patient characteristics by hospital safety-net burden group. We calculated quality metric concordance rates for each of the 20 quality metrics, defined as the fraction of patients receiving concordant care stratified by safety-net status. Keeping consistent with Medicare’s pay-for-performance reimbursement models, we risk-adjusted our quality concordance rates to remove the hospital-specific imbalance of patient age, gender, comorbidity, and differences in stage at presentation^4,19-22^.

We conducted subset analyses among high-burden hospitals to determine how quality metric concordance rates vary by different hospital characteristics. This subset analysis evaluated the total hospital concordance level with all metrics. Hospital characteristics assessed included the hospital safety-net burden (fraction of uninsured or Medicaid patients), as well as the proportion of minority patients the hospital serves (non-White or Hispanic patients), and income and education level of patients. This subset analysis consisted of a linear regression treating each hospital as an independent observation, weighting the hospitals by their patient volume.

Statistical analyses were conducted with SAS (Version 9.4; SAS Institute Inc., Cary, NC), and regression figures were generated with R (Version 3.6.1; The R Foundation for Statistical Computing, Vienna, Austria). Statistical tests were two-sided, and p-values < 0.05 were considered statistically significant.

## Results

### Hospital and patient characteristics

Our study cohort included 1,703,865 cancer patients treated at 1,344 hospitals. This included 326 low-burden hospitals (lowest quartile of uninsured or Medicaid patients), 668 middle-burden hospitals, and 350 high-burden hospitals (highest quartile of uninsured or Medicaid patients). High-burden hospitals treated a median of 13.4% uninsured or Medicaid patients, compared to 7.7% for middle-burden hospitals, and 3.9% for low-burden hospitals. In general, higher-burden hospitals were more likely to be academic and located in the Midwest and West, compared to middle- and low-burden hospitals. High-burden hospitals were also more likely to treat black, Hispanic, younger, male, low education, low income, and advanced stage patients compared to middle- and low-burden hospitals. See **Table 2** for full descriptive characteristics of the study subjects stratified by hospital.

### Concordance with quality of care measures

The general level of quality metric concordance across the whole study population varied substantially by the individual quality metric, ranging from a low of 37.5% with appropriate lymph node removal in surgery for lung cancer to a high of 91.0% with appropriate radiation after breast conserving surgery (**Table 3**). Quality metrics involving surgery (appropriate surgery or lymph node removal) had the lowest concordance rates (mean 67.7%), followed by radiation alone (78.2%), chemotherapy or hormonal therapy alone (79.1%), and combination chemotherapy/radiation (81.1%). High-burden hospitals were significantly less likely to meet recommendations for 13 of 20 quality measures (65%) compared to low-burden hospitals (**Table 3**). Among the underperforming low-burden hospitals, the quality gap in concordance levels between high- and low-burden hospitals ranged from 2.1% to 11.1%. High-burden hospitals underperformed low-burden hospitals in 4 of 8 quality measures related to surgery (50%), in 5 of 5 quality measures related to chemotherapy or hormonal therapy alone (100%), in 3 of 4 quality measures related to radiotherapy alone (75%), and in 1 of 3 quality measures related to combination chemoradiotherapy (33.3%). Of note, high-burden hospitals underperformed low-burden hospitals in 5 of 5 “accountability” quality measures (100%) which reflect measures backed by the highest levels of evidence. In 1 of 20 quality measures, the high-burden hospitals outperformed low burden hospitals – this specific quality metric represented overuse of surgery in lung cancer, with low-burden hospitals more likely to overuse surgery compared to high-burden hospitals.

**Table 3.**
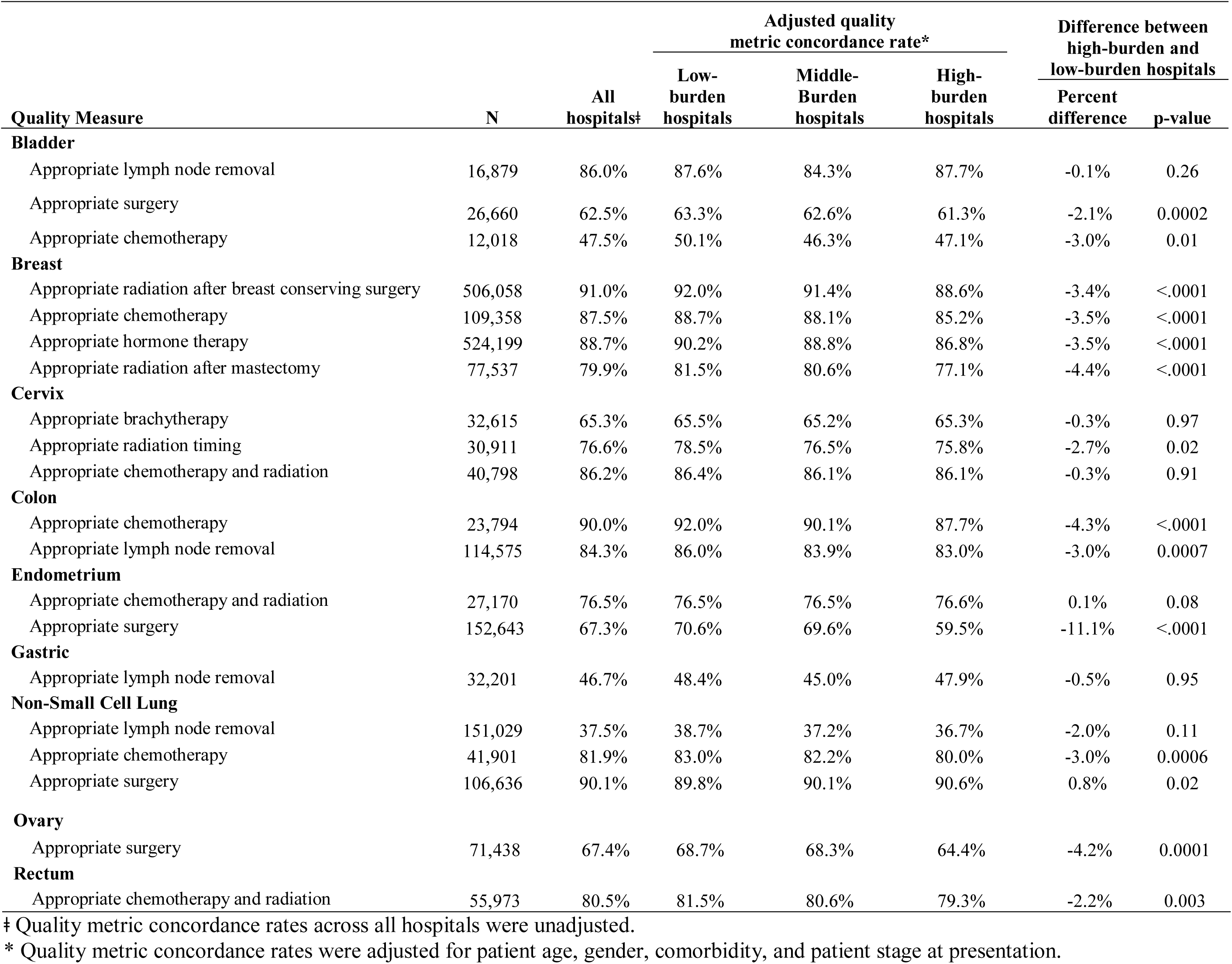
Concordance with Quality Measures by Safety-Net Burden.

| Quality Measure | N | All hospitals† | Adjusted quality metric concordance rate* |  |  | Difference between high-burden and low-burden hospitals |  |
| --- | --- | --- | --- | --- | --- | --- | --- |
|  |  |  | Low-burden hospitals | Middle-Burden hospitals | High-burden hospitals | Percent difference | p-value |
| Bladder |  |  |  |  |  |  |  |
| Appropriate lymph node removal | 16,879 | 86.0% | 87.6% | 84.3% | 87.7% | -0.1% | 0.26 |
| Appropriate surgery | 26,660 | 62.5% | 63.3% | 62.6% | 61.3% | -2.1% | 0.0002 |
| Appropriate chemotherapy | 12,018 | 47.5% | 50.1% | 46.3% | 47.1% | -3.0% | 0.01 |
| Breast |  |  |  |  |  |  |  |
| Appropriate radiation after breast conserving surgery | 506,058 | 91.0% | 92.0% | 91.4% | 88.6% | -3.4% | <.0001 |
| Appropriate chemotherapy | 109,358 | 87.5% | 88.7% | 88.1% | 85.2% | -3.5% | <.0001 |
| Appropriate hormone therapy | 524,199 | 88.7% | 90.2% | 88.8% | 86.8% | -3.5% | <.0001 |
| Appropriate radiation after mastectomy | 77,537 | 79.9% | 81.5% | 80.6% | 77.1% | -4.4% | <.0001 |
| Cervix |  |  |  |  |  |  |  |
| Appropriate brachytherapy | 32,615 | 65.3% | 65.5% | 65.2% | 65.3% | -0.3% | 0.97 |
| Appropriate radiation timing | 30,911 | 76.6% | 78.5% | 76.5% | 75.8% | -2.7% | 0.02 |
| Appropriate chemotherapy and radiation | 40,798 | 86.2% | 86.4% | 86.1% | 86.1% | -0.3% | 0.91 |
| Colon |  |  |  |  |  |  |  |
| Appropriate chemotherapy | 23,794 | 90.0% | 92.0% | 90.1% | 87.7% | -4.3% | <.0001 |
| Appropriate lymph node removal | 114,575 | 84.3% | 86.0% | 83.9% | 83.0% | -3.0% | 0.0007 |
| Endometrium |  |  |  |  |  |  |  |
| Appropriate chemotherapy and radiation | 27,170 | 76.5% | 76.5% | 76.5% | 76.6% | 0.1% | 0.08 |
| Appropriate surgery | 152,643 | 67.3% | 70.6% | 69.6% | 59.5% | -11.1% | <.0001 |
| Gastric |  |  |  |  |  |  |  |
| Appropriate lymph node removal | 32,201 | 46.7% | 48.4% | 45.0% | 47.9% | -0.5% | 0.95 |
| Non-Small Cell Lung |  |  |  |  |  |  |  |
| Appropriate lymph node removal | 151,029 | 37.5% | 38.7% | 37.2% | 36.7% | -2.0% | 0.11 |
| Appropriate chemotherapy | 41,901 | 81.9% | 83.0% | 82.2% | 80.0% | -3.0% | 0.0006 |
| Appropriate surgery | 106,636 | 90.1% | 89.8% | 90.1% | 90.6% | 0.8% | 0.02 |
| Ovary |  |  |  |  |  |  |  |
| Appropriate surgery | 71,438 | 67.4% | 68.7% | 68.3% | 64.4% | -4.2% | 0.0001 |
| Rectum |  |  |  |  |  |  |  |
| Appropriate chemotherapy and radiation | 55,973 | 80.5% | 81.5% | 80.6% | 79.3% | -2.2% | 0.003 |

‡ Quality metric concordance rates across all hospitals were unadjusted.
\* Quality metric concordance rates were adjusted for patient age, gender, comorbidity, and patient stage at presentation.

### Variation in quality among high-burden hospitals

Among the 350 high-burden hospitals, we found that overall hospital guideline concordance rates varied by hospital characteristics (**Figure 1**). Hospitals caring for larger fractions of uninsured or Medicaid patients (**Figure 1A**), minority serving hospitals (**Figure 1B**), and hospitals caring for less educated patients (**Figure 1D**) all had significantly lower levels of guideline concordance with quality measures (all p<0.001). We found no correlation between income level of patients and guideline concordance rates (**Figure 1C**; p=0.43).

**Figure 1.**
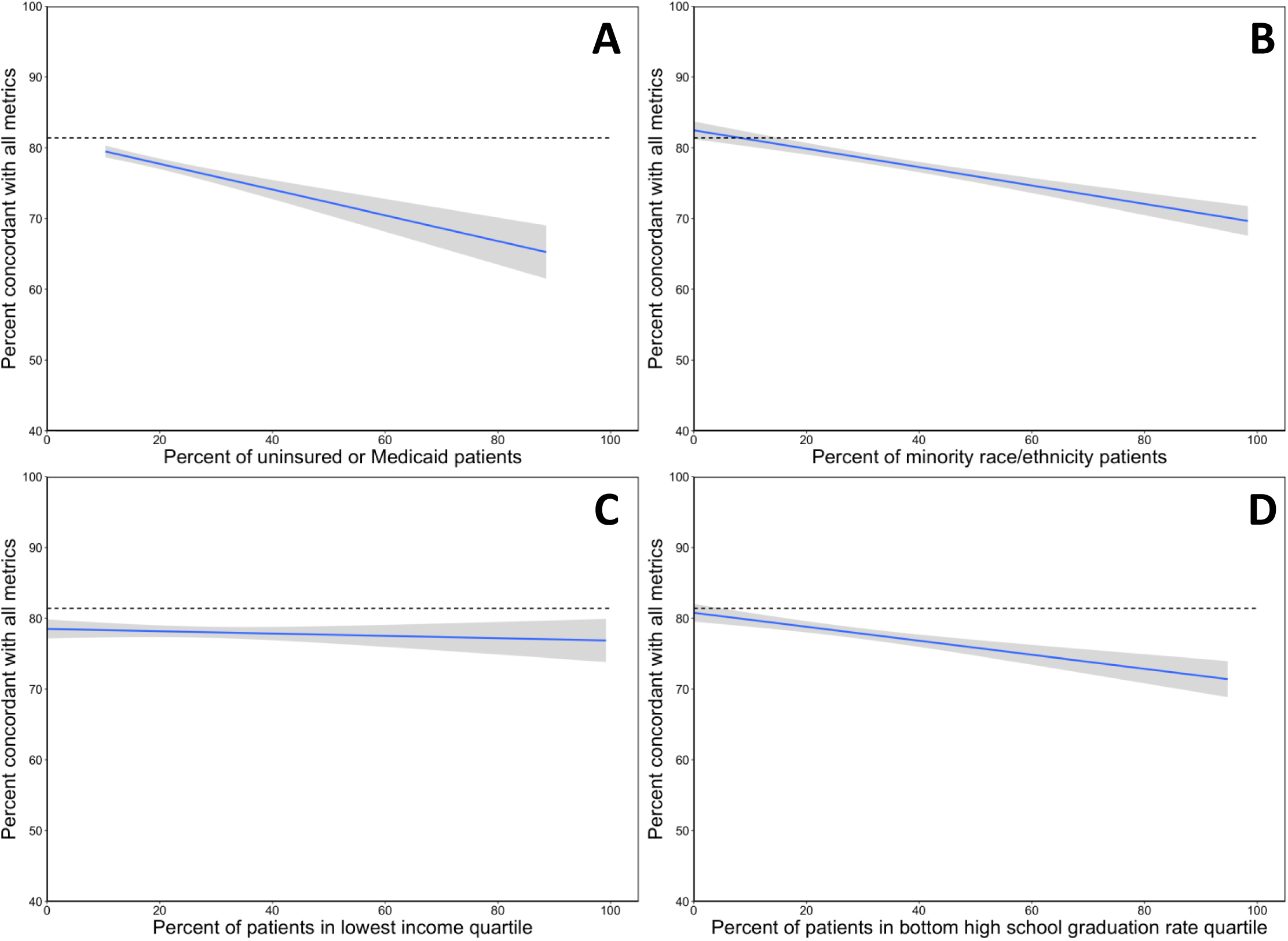
Correlation between hospital characteristics and concordance with quality metrics. This figure represents an analysis among the high-burden hospitals (hospitals caring for highest quartile of uninsured or Medicaid patients) comparing hospital-wide characteristics to concordance with quality of care metrics. The solid line represents the relationship between hospital characteristic and concordance level, and the shaded region represents the 95% confidence band. The dotted line represents the average quality metric concordance level among low- and medium-burden hospitals (81.6%).

## Discussion

This current study with over 1.7 million cancer patients demonstrates that hospitals caring for a disproportionate number of uninsured or Medicaid patients have a lower likelihood of providing care concordant with standard quality measures. While our study represents the first to evaluate concordance with quality of care measures, others have studied patterns of care and outcomes at safety-net hospitals. Themes that arise among patients treated at high-burden hospitals include longer hospital stays, higher rates of operative complications, increased mortality, and increased costs of care^18,23^,24. Our study supports this existing literature, and overall research demonstrates a consistent association between high-burden hospitals and decreased quality of care.

The association between hospital safety-net status and decreased quality of care represents an important point worth further discussion. One must consider that this association between hospital safety-net status and patient outcomes does not imply causality. Safety-net hospitals by definition provide care for poor and disadvantaged patients. This current study supports prior research^25,26^, which demonstrates that hospitals with a higher safety-net burden care for a higher fraction of racial and ethnic minorities, with decreased income, lower levels of education, and more advanced stage cancer on presentation. These at-risk patients may have economic, social, or health care access-related barriers which directly influence the quality and timeliness of care. A substantial body of literature demonstrates that these social determinants of health independently influence patterns of healthcare delivery^27,28.^ Hospitals should strive to provide equal care to all patients irrespective of their sociodemographic backgrounds; however, providing high quality timely care can prove challenging with our most underserved patients.

Healthcare is rapidly transitioning from a fee-for-service reimbursement model to pay-for-performance reimbursement, which strives to reward hospitals and providers for delivering high-quality healthcare. While pay-for-performance reimbursement models may appeal to both payers and stakeholders, one must consider the impact these alternative reimbursement policies will have on health disparities. In general, quality of care measurements represent a central feature in value-based reimbursement models. For example, the Oncology Care Model, an alternative payment model currently being piloted by CMS, includes four clinical care quality measures^29^. Three of these four quality measures were evaluated in this current study, and high-burden hospitals underperformed low-burden hospitals in all three (chemotherapy in colon cancer, chemotherapy in breast cancer, hormone therapy in breast cancer). Similar to CMS, private insurers are also piloting alternative payment models^30-32^. This current study points to a concerning reality that moving to a pay-for-performance reimbursement paradigm in oncology may further decrease payments to hospitals caring for underserved and minority patients.

This current study highlights another area of concern in that among safety-net hospitals, those with the lowest concordance with quality metrics cared for the highest fraction of underinsured, minority, and uneducated patients. This suggests that hospitals caring for the most at-risk patients may incur the highest financial penalties under pay-for-performance reimbursement models. Our finding of safety-net hospital vulnerability with oncology reimbursement supports research in other areas in healthcare demonstrating that value-based payments can exacerbate health disparities^33^. A recent study by Chen et al., evaluated the Medicare Physician Value-Based Payment Modifier Program, initiated in 2015, which provides bonuses to practices based on quality and costs of care. This study found that practices caring for more socially at risk patients had lower quality scores, fewer bonuses and more penalties^34^. This so-called “reverse Robin Hood” effect takes money from underserved hospitals (in the form of penalties) and gives it to wealthier hospitals (in the form of bonuses)^35^.

Ideally, payment models should strive to define fair, equitable, and non-discriminatory quality measures upon which to base reimbursement. However, identifying good quality measures independent of race or socioeconomic status in oncology, and in medicine in general, can prove difficult^36^. Regardless, value-based reimbursement models should consider actionable solutions to avoid expanding health disparities. Adjusting quality scores for a patient’s race and socioeconomic status represents one potential solution. While this approach might help reduce the reimbursement gap it also in theory sends a message that poor quality of care is acceptable at hospitals serving underserved patients. Another pay-for-performance approach to minimize disparities would include comparing “apples to apples”^33^ – or in other words, compare hospitals based on their patient-risk profile. This would include comparing performance for safety-net hospitals to other safety-net hospitals, which research demonstrates has the potential to substantially reduce disparities in payments across organizations^37^. Regardless of the solution, researchers and policymakers should continue to focus on the complex intersection of reimbursement policy and health equity to help optimize patient care for the underserved.

This retrospective observational study has potential limitations worth considering. First, the NCDB does not capture information on patient variables including performance status, body habitus, smoking status, and other risk factors which may help explain the differences in concordance to quality of care measures. However, it would be unlikely for these risk factors to factor into any risk-adjustment protocol given the difficulty of measuring these data points at a national level. Overall, the data recorded within NCDB reflect the data used in current quality measures in oncology^29^. Another potential limitation refers to generalizability. While the NCDB includes over 70% of incident cancer cases in the US, this only includes Commission on Cancer accredited facilities. Given the barriers to accreditation and costs associated with data collection, it is likely that under-resourced safety-net hospitals could be underrepresented in our study population. However, including these more under-resourced facilities in our study may in fact widen the gap between high-burden and low-burden hospitals, though more research is needed to address this question. Lastly, our assessment of guideline concordance depends on data captured by individual hospital registrars. Any inaccuracies in data collection would result in misclassification of our endpoints, and could influence our results. However, individual facilities undergo credentialing and internal quality assurance procedures^38^ to assure high quality of data collection. Despite this internal quality assurance, misclassification could add bias to the study findings. Though one must consider that the data captured by registrars is the same data used to determine hospital performance metrics, therefore this misclassifications would translate into bonuses or penalties under a pay-for-performance reimbursement model.

Despite these limitations, our results highlight a concerning pattern of diminished quality of care among hospitals caring for disproportionate numbers of uninsured or Medicaid patients. These differences in quality of care in part reflect the social and economic challenges faced by our underserved patients. As we move towards pay-for-performance healthcare, these quality gaps will translate to reimbursement gaps, which could in fact exacerbate health disparities among our underserved population. Value-based reimbursement represents the future of healthcare; however, policymakers must remain cognizant of the impact that reimbursement policy has on our at-risk population.

## Data Availability

All data referred to in the manuscript is publicly available through the National Cancer Database (NCDB).

## REFERENCES

1. New England Journal of Medicine Catalyst: What Is Pay for Performance in Healthcare? New England Journal of Medicine Catalyst, Massachusettes Medical Society, 2018

2. Kuhmerker K, Hartman T: Pay-For-Performance In State Medicaid Programs: A Survey of State Medicaid Directors and Programs. The Commonwealth Fund, 2007

3. Rosenthal MB, Landon BE, Normand S-LT, et al: Pay for Performance in Commercial HMOs. New England Journal of Medicine 355:1895–1902, 2006

4. Suter LG, Vellanky S, Li S-X, et al: Medicare Hospital Quality Chartbook 2012: Performance Report on Outcome Measures in Yale New Haven Health Services Corporation Center for Outcomes Research and Evaluation (ed), Centers for Medicare & Medicaid Services, 2012

5. Irons TG, Moore KS: The importance of health insurance and the safety net in rural communities. N C Med J 76:50–3, 2015

6. Farkas DT, Greenbaum A, Singhal V, et al: Effect of insurance status on the stage of breast and colorectal cancers in a safety-net hospital. J Oncol Pract 8:16s–21s, 2012

7. Werner RM, Goldman LE, Dudley RA: Comparison of change in quality of care between safety-net and non-safety-net hospitals. JAMA 299:2180–7, 2008

8. McAlearney AS, Murray K, Sieck C, et al: The Challenge of Improving Breast Cancer Care Coordination in Safety-net Hospitals: Barriers, Facilitators, and Opportunities. Med Care 54:147–54, 2016

9. Figueroa JF, Joynt KE, Zhou X, et al: Safety-net Hospitals Face More Barriers Yet Use Fewer Strategies to Reduce Readmissions. Med Care 55:229–235, 2017

10. Andrulis DP, Siddiqui NJ: Health Reform Holds Both Risks And Rewards For Safety-Net Providers And Racially And Ethnically Diverse Patients. Health Affairs 30:1830–1836, 2011

11. Gaskin DJ, Hadley J: Population characteristics of markets of safety-net and non-safety-net hospitals. J Urban Health 76:351–70, 1999

12. Gilman M, Adams EK, Hockenberry JM, et al: Safety-Net Hospitals More Likely Than Other Hospitals To Fare Poorly Under Medicare’s Value-Based Purchasing. Health Affairs 34:398–405, 2015

13. Joynt KE, Jha AK: Characteristics of Hospitals Receiving Penalties Under the Hospital Readmissions Reduction Program. JAMA 309:342–343, 2013

14. Commission on Cancer: About the Commission on Cancer, American College of Surgeons, 2019

15. Shulman LN, McCabe R, Gay G, et al: Building Data Infrastructure to Evaluate and Improve Quality: The National Cancer Data Base and the Commission on Cancer’s Quality Improvement Programs. J Oncol Pract 11:209–12, 2015

16. American College of Surgeons: CoC Quality of Care Measures, American College of Surgeons, 2019

17. Lewin ME, Altman S: Americas’s Health Care Safety Net: Intact but Endangered, Americas’s Health Care Safety Net: Intact but Endangered. Washington (DC), Institute of Medicine (US) Committee on the Changing Market, Managed Care, and the Future Viability of Safety Net Providers, 2000

18. Hoehn RS, Wima K, Vestal MA, et al: Effect of Hospital Safety-Net Burden on Cost and Outcomes After Surgery. JAMA Surg 151:120–8, 2016

19. Centers for Medicare & Medicaid Services: Hospital-Acquired Condition Reduction Program: Hospital-Specific Report User Guide Fiscal Year 2020, 2019

20. Centers for Medicare & Medicaid Services: Hospital-Specific Report User Guide: Hospital Readmissions Reduction Program Fiscal Year 2020 2019

21. Centers for Medicare & Medicaid Services: Hospital Value-Based Purchasing (VBP) Program: Hospital-Specific Report User Guide for the Mortality and Complication Measures Fiscal Year 2020, 2019

22. Centers for Medicare & Medicaid Services Innovation Center: Oncology Care Model Overview, 2019

23. Sarkar R, Guss ZD, Lopez C, et al: Impact of hospital safety-net burden on oncology patterns of care and outcomes. Journal of Clinical Oncology 36:6567–6567, 2018

24. Wakeam E, Hevelone ND, Maine R, et al: Failure to rescue in safety-net hospitals: availability of hospital resources and differences in performance. JAMA Surg 149:229–35, 2014

25. Rhoads KF, Ackerson LK, Jha AK, et al: Quality of colon cancer outcomes in hospitals with a high percentage of Medicaid patients. J Am Coll Surg 207:197–204, 2008

26. Porten SP, Richardson DA, Odisho AY, et al: Disproportionate presentation of high risk prostate cancer in a safety net health system. J Urol 184:1931–6, 2010

27. Braveman P, Gottlieb L: The social determinants of health: it’s time to consider the causes of the causes. Public Health Rep 129 Suppl 2:19–31, 2014

28. Daniel H, Bornstein SS, Kane GC, et al: Addressing Social Determinants to Improve Patient Care and Promote Health Equity: An American College of Physicians Position Paper. Ann Intern Med 168:577–578, 2018

29. Centers for Medicare and Medicaid Services: Oncology Care Model, 2019

30. Association of Northern California Oncologists: An Introduction to Innovent Oncology: A Service of US Oncology, 2019

31. Newcomer LN, Gould B, Page RD, et al: Changing Physician Incentives for Affordable, Quality Cancer Care: Results of an Episode Payment Model. Journal of Oncology Practice 10:322–326, 2014

32. Penne J: MD Anderson, UnitedHealthcare Launch New Cancer Care Payment Model, MD Anderson Cancer Center, 2014

33. Rubin R: How Value-Based Medicare Payments Exacerbate Health Care Disparities. JAMA 319:968–970, 2018

34. Chen LM, Epstein AM, Orav EJ, et al: Association of Practice-Level Social and Medical Risk With Performance in the Medicare Physician Value-Based Payment Modifier Program. JAMA 318:453–461, 2017

35. Frakt AB, Jha AK: Face the Facts: We Need to Change the Way We Do Pay for Performance. Ann Intern Med 168:291–292, 2018

36. Valuck T, Blaisdell D, Dugan D, et al: Improving Oncology Quality Measurement in Accountable Care, 2017

37. Damberg CL, Elliott MN, Ewing BA: Pay-for-performance schemes that use patient and provider categories would reduce payment disparities. Health Aff (Millwood) 34:134–42, 2015

38. Commission on Cancer: Cancer Program Standards: Ensuring Patient-Centered Care 2016 Edition, American College of Surgeons, 2016

